# Gait event prediction from surface electromyography in parkinsonian patients

**DOI:** 10.1101/2023.01.13.22282375

**Authors:** Stefan Haufe, Ioannis U. Isaias, Franziska Pellegrini, Chiara Palmisano

**Author notes:** (C.P.); (I.U.I.).

## Abstract

Gait disturbances are common manifestations of Parkinson’s disease (PD), with unmet therapeutic needs. Inertial measurement units (IMU) are capable of monitoring gait, but they lack neurophysiological information that may be crucial for studying gait disturbances in these patients. Here, we present a machine-learning approach to approximate IMU angular velocity profiles, and subsequently gait events from electromyographic (EMG) channels. We recorded six parkinsonian patients while walking for at least three minutes. Patient-agnostic regression models were trained on temporally-embedded EMG time series of different combinations of up to five leg muscles bilaterally (i.e., tibialis anterior, soleus, gastrocnemius medialis, gastrocnemius lateralis, and vastus lateralis). Gait events could be detected with high temporal precision (median displacement <50 msec), low numbers of missed events (<2%), and next to no false positive event detections (<0.1%). Swing and stance phases could thus be determined with high fidelity (median F1 score ∼0.9). Interestingly, the best performance was obtained using as few as two EMG probes placed on the left and right vastus lateralis. Our results demonstrate the practical utility of the proposed EMG-based system for gait event prediction while allowing the simultaneous acquisition of an electromyographic signal. This gait analysis approach has the potential to make additional measurement devices such as IMU and force plates less essential, and thereby to reduce financial and preparation overheads and discomfort factors in gait studies.

## 1. Introduction

Gait and balance disturbances are common and important clinical manifestations of Parkinson’s disease (PD), leading to mobility impairment and falls [1]. Current treatments (pharmacological and deep brain stimulation, DBS) provide only partial benefits in gait derangements in PD, with a wide variability in outcomes [2–5].

Despite detailed testing, specific factors that are critical to predict locomotor deterioration in PD remain elusive [6–9]. Besides the subtle onset and clinical heterogeneity [10], technical limitations have hampered the timely and direct recording of supraspinal locomotor derangements in these patients. Only recently have advances in portable electroencephalography systems [11,12] and new DBS devices capable of on-demand recording from the chronically-implanted electrodes (e.g., Activa PC+S and Percept PC [Medtronic PLC] or AlphaDBS [Newronika Srl]) [13–15] enabled recording of ongoing brain activity during actual gait in PD [16–18].

Precise assessment of gait dynamics should account for its context dependency. New study setups employing fully-immersive virtual reality (VR) or augmented reality allow gait assessment (with optoelectronic systems, force plates, etc.) in environments that deliver patient-specific triggers of gait impairment (e.g., [19]). These setups could facilitate the identification of biomarkers for the fine-tuning of therapy delivery, e.g., adaptive DBS programming and so-called VR Exposure Therapy [20].

An open challenge is the continuous monitoring of gait parameters in laboratory as well as real-world environments. Technically, parameters such as timings of heel-strike and toe-off events, which define swing and stance phases and provide valuable information about cadence patterns, etc., can be assessed with optoelectronic systems and force plates. Both systems, however, are expensive, require qualified personnel and do not offer monitoring in ecological settings. Video-based analyses of gait have been proposed as well [21], although it is unclear whether these would reach the required precision to identify individual events within a gait cycle, especially for clinical applications and in ecological settings. Wearable motion sensors such as inertial measurement units (IMU) are another viable option to capture gait events in natural environments with high temporal accuracy [22,23]. However, they do not contain further neurophysiological information that may be crucial to understand and predict gait derangements [24]. Surface electromyography (EMG) provides the missing link between neural signals and kinematics that enables comprehensive characterization of pathological gait. EMG measurements have been used to predict lower limb motion in advance [25,26] for real-time control of a prosthesis [27–29] or adaptive DBS devices [16–18,25,26,30,31]. EMG profiles of the gait cycle have also been shown to anticipate specific gait derangements in PD such as freezing of gait [32], a sudden episodic inability to produce effective stepping despite the intention to walk. The combined use of IMU and EMG signals would enable description of the motor actions and intentions underlying gait kinematic features and alterations.

However, some practical limitations should be considered when applying additional sensors on severely ill patients, especially when performing recordings after suspension of medications. For example, in patients with PD, overnight suspension of dopaminergic drugs is fundamental to evoke and study PD-related symptoms, but greatly reduces the time window available for experimental recordings. Limiting the preparation period by limiting the number of sensors may help considerably in this regard. Also, an excessive number of sensors may alter the natural behavior of subjects, undermining the advantages of working in ecological environments. Another crucial aspect is the cost of multiple sets of sensors. Considering that probes comprising both IMU and EMG are generally more expensive than standalone solutions, the need for IMU and EMG in the fine-grained evaluation of gait may be a limiting factor for many laboratories and applications in clinical routine. The use of multiple devices may also not be practical in clinical routine, as synchronization or different recording software may be needed.

Considering this, the development of novel technologies that can extract multiple types of signals from the same set of sensors is highly desirable. While the same kinematics can be produced by different muscular patterns, lower limb kinematics can be inferred by analysis of the EMG [33]. The idea of detecting gait events directly from EMG signals, circumventing additional IMU or force plates, is gaining traction [34–38]. Ziegler and colleagues [39] report high accuracy in classifying the stance and swing phases during human gait based on EMG recordings. They first extracted a weighted signal difference that exploits the difference of the EMG activity between corresponding muscles of the two legs, and then trained a support vector machine to classify gait phases. Using a deep learning approach, Morbidoni and colleagues [35] were also able to classify stance and swing phases and predict foot-floor contacts in natural walking conditions in healthy subjects. Other studies showed similar results in learned and unlearned subjects [37], and using intra-subject training only [34]. This would not only simplify future recording setups but also permit the re-analysis of EMG datasets recorded without IMU or in cases of data loss due to technical problems with the IMU. This second scenario is particularly problematic when recordings cannot be repeated due to the patient’s clinical condition. Additionally, the extraction and prediction of gait events from lower limb EMG activity is of fundamental importance for the development of an EMG-driven prosthesis, where predicting the subsequent gait phase from muscular signals increases prosthesis efficiency and responsiveness [33]. To the best of our knowledge, no approach has yet aimed to reconstruct complete time series encoding gait-related activity such as IMU traces instead of discrete gait events. Moreover, we are not aware of any approach that has been tested on unmedicated PD patients during long periods of continuous walking.

In the present study, we explore the possibility of identifying fundamental gait events from surface EMG in parkinsonian patients using a machine-learning approach. Compared to previous studies, we did not frame the problem as one of detection (i.e., to identify the timings of a fixed set of events) or classification (i.e., to segment the data into contiguous gait phases). Instead, we used an innovative regression approach to approximate continuous angular velocity profiles as measured by IMU. We consider this approach strictly more powerful and flexible than previous approaches, as access to the predicted IMU time series allows us not only to extract predetermined types of gait events but also biomechanical quantities such as joint angular velocity and further parameters on which our model has not been trained. Our study is further set apart from published work in that we focus on a clinical cohort rather than healthy participants. To our knowledge, our study is the first to demonstrate the feasibility of accurate gait parameter estimation from EMG in such a population. Remarkably, our approach accounts for the substantial across-patient variability observed in gait patterns of clinical populations, allowing it to be applied without any patient-specific calibration.

## 2. Materials and Methods

### 2.1. Participants

We recruited six patients with idiopathic PD according to the UK Brain Bank criteria who did not suffer from any other disease, including cognitive decline (i.e., Mini-Mental State Examination score >27), vestibular disorders, and orthopedic impairments that could interfere with walking. An additional inclusion criterion for this preliminary study was the ability of the patient to walk continuously and without assistance for at least three minutes. Disease severity was evaluated using the MDS-Unified Parkinson’s Disease Rating Scale motor part (UPDRS-III) and the stage of the disease using the Hoehn and Yahr (H&Y) scale. Using items 3.15–3.17 of the UPDRS-III (hands and feet), a sum rest tremor sub-score was created for the right and left side separately. Similarly, a sum bradykinesia-rigidity score (items 3.3–3.8) was obtained for each side.

Demographic and clinical features are listed in Table 1. The study was approved by the Ethics Committee of the University of Würzburg and conformed to the declaration of Helsinki. All patients gave their written informed consent to participation.

**Table 1.** Demographic and clinical features. Abbreviations: Hoehn and Yahr stage (H&Y); Levodopa equivalent daily dose (LEDD); Unified Parkinson’s Disease Rating Scale motor part (UPDRS-III); Meds-off: practical medication-off state, i.e., overnight withdrawal (>12 h) of all dopaminergic drugs; Meds-on: medications-on state 30-60 min after receiving 1 to 1.5 times the levodopa-equivalent of the morning dose. UPDRS-III is presented as total score/tremor sub-score left/tremor sub-score right/bradykinesia-rigidity sub-score left/bradykinesia-rigidity sub-score right.

|  | Gender | Age, years | Age<br>at onset, years | LEDD, mg | UPDRS-III<br>meds-off | UPDRS-III<br>meds-on | H&Y |
| --- | --- | --- | --- | --- | --- | --- | --- |
| WP1 | M | 46-50 | 36-40 | 1167 | 50/2/4/14/11 | 15/1/0/3/4 | 3 |
| WP2 | M | 56-60 | 45-50 | 900 | 28/3/7/4/9 | 5/0/0/0/4 | 2 |
| WP3 | F | 56-60 | 51-55 | 362 | 18/2/0/2/8 | 11/1/0/1/7 | 1 |
| WP4 | F | 51-55 | 45-50 | 640 | 9/0/0/6/2 | 5/0/0/4/1 | 1 |
| WP5 | M | 61-65 | 51-55 | 610 | 12/0/0/2/8 | 5/0/0/0/4 | 2 |
| WP6 | M | 61-65 | 56-60 | 610 | 30/0/1/5/13 | 21/0/0/2/8 | 2 |

### 2.2. Experimental setup and procedure

Patients were investigated in a practical medication-off state, i.e., on the morning after overnight withdrawal (>12 h) of all dopaminergic drugs (meds-off). Kinematic data were recorded using two IMU (Opal, APDM), with a sampling rate of 128 Hz, placed bilaterally on the outer anklebones. Each sensor was placed with its vertical axis aligned to the tibial anatomic axis. Surface leg muscle activity as measured by 10 EMG probes (FREEEMG, BTS) was recorded bilaterally from the tibialis anterior (Ta), soleus (S), gastrocnemius medialis (Gm), gastrocnemius lateralis (Gl), and vastus lateralis (Vl), at a sampling rate of 1000 Hz. Two transistor-transistor logic signals (TTL) were provided at the beginning and end of each trial to both EMG and IMU devices to allow data synchronization. Patients started walking barefoot after a verbal signal at their self-selected speed along a large ellipsoidal path of about 60 m length. We recorded between three to six trials (243 ±71 sec duration) of unperturbed, steady-state, overground walking according to the clinical condition of each subject. Overall, 26 walking trials with a total duration of 105 min were obtained.

### 2.3. Selection of EMG channels

We focused on the muscles of the lower leg, which are highly involved during the gait cycle. The Ta and S are distal monoarticular muscles with distinct and synergistic contributions to human gait [40]. According to [41], they are the most active muscles during gait and display the lowest inter-subject variability. We therefore hypothesized that models based on bilateral pairs of these muscles may be particularly suitable and potentially sufficient for predicting gait-related angular velocity profiles. The gastrocnemius muscle (biarticular) was added for a comprehensive evaluation of the triceps surae. Note that, since medial and lateral gastrocnemii fulfill somewhat independent roles [42,43], both were added. Given the knee flexor activity of the gastrocnemius muscle, we then positioned the last available probe on the Vl, a major (monoarticular) knee extensor muscle.

Models based on different muscle combinations were compared to the model including all five pairs of muscles. We were interested in identifying minimal subsets of EMG probes that would enable accurate IMU reconstruction. Thus, we further exhaustively tested all possible 2^5^-1 = 31 sets containing between one and five pairs of distinct muscles. Note that all considered models included either none or both the left and right EMG signals for each studied muscle. Thus, all models comprised an even number of muscles between two and ten.

### 2.4. Data preprocessing

EMG data were bandpass-filtered, rectified, and down-sampled to 200 Hz. IMU traces were up-sampled to 200 Hz using nearest-neighbor interpolation. IMU and EMG data were aligned to the rising edge of the first TTL signal for synchronization. A number of preprocessing steps were devised to facilitate the prediction of angular velocity traces from EMG data. To smooth out local extrema occurring due to noise, IMU data were processed with a moving-median filter with 100 msec window length, followed by a moving-mean filter with 40 msec window length. To achieve a similar degree of smoothness, EMG data were processed with a moving-median filter with 200 msec window length, followed by a moving-mean filter with 40 msec window length. All moving filters were centered. As a simple high-pass filter, the minimum in a moving window of 10 sec length was subtracted from the EMG data. To standardize scales across patients, EMG activation time courses were further normalized by subtracting the first percentile and dividing by the 95^th^ percentile. Percentiles were estimated separately for each recording. Each recording was cropped to the exact onand offsets of the walking period.

### 2.5. Extraction of biomechanical parameters

Swing peak velocity (SWP), heel contact (HC), and toe-off (TO) events were extracted from the angular velocity profiles measured with respect to the medio-lateral axis by the IMU (see [44] and [45] for an extensive description on gait event detection in IMU data). This was done separately for the left and right IMU sensor as follows. First, SWP events were identified as local maxima with at least 150°/sec peak height and 0.7 sec inter-peak distance. Two consecutive SWP events defined one gait cycle. Next, local minima within each cycle were used to define corresponding HC and TO events. The HC event was defined as the earliest local minimum occurring in the sub-interval between 10% and 45% of the cycle. If no local minimum could be found, the global minimum within that sub-interval was used. Similarly, the TO events were defined as the latest local minimum occurring in the sub-interval between 55% and 90% of the cycle. Again, if no local minimum could be found, the global minimum within that sub-interval was used. At random, events extracted by the described algorithm were checked by an expert (C.P.) and were in agreement with a manual determination based on the same IMU data. The procedure was used to define “ground-truth” gait events from recorded IMU data, as well as approximate event timings derived from reconstructed angular velocity time series based on EMG activity (see below).

### 2.6. Prediction of angular velocity profiles from EMG

We used multiple linear regression to approximate the angular velocity with respect to the medio-lateral axis of the left and right ankle from the combined activation traces of multiple muscles within a window around the prediction point. The regression coefficients were fitted to minimize the mean-squared error between measured and approximated IMU traces on training data, consisting of pairs of IMU and EMG activity traces. To enable the prediction model to utilize the temporal dynamics of the EMG channels around the prediction time point, a temporal embedding of the EMG time series was performed. To this end, each selected EMG channel was complemented by temporally-shifted versions 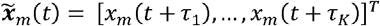, m = 1, …, *M*, where *x*_*m*_ (*t*+ *τ* was the activity of the m-th EMG sensor at time *t*+ *τ*. Here, we used *K*=21 equally spaced shifts, ranging from *τ*_1_ = -500 msec to *τ* = +500 msec in steps of 50 msec. Thus, the prediction of the IMU signal at time *t* was based on EMG information within a window around t of one second length. The relation between the embedded signal of all *M* EMG sensors, 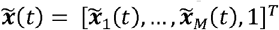 (including an offset term) and the angular velocity *y*(*t*) (either at the left or right ankle) was assumed to be linear according to the model 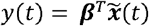. The (*K* • *M*+ 1) -dimensional coefficient vector 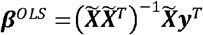 was estimated using ordinary least-squares (OLS) regression, where 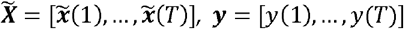, and T denoted the number of available paired measurements of EMG and IMU activity in the training set. Using the fitted model, EMG-based IMU predictions were obtained as 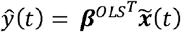.

### 2.7. Performance analysis

Models were evaluated exclusively on holdout data using leave-one-patient-out cross-validation. Models were fitted on the concatenated trials of all but one patient (the training data) and were evaluated on all trials of the held-out (test) patient, where each patient served as the hold-out patient once. This evaluation scheme provided an unbiased assessment of the prediction performance. Model predictions 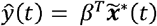 were obtained by multiplying the coefficient vector *β* estimated on the training data to embedded EMG data 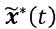 from each trial of the test patient.

Predicted and ground-truth data were compared on a per-trial and leg basis using the Pearson correlation coefficient (r). Gait events (SWP, HC, and TO) were extracted from the predicted IMU time series as described in Section 2.5. Separately for each leg, ground-truth and predicted HC and TO events were used to divide each trial into alternating segments representing the swing and stance phases of the gait cycle. The resulting binary time series were compared using the F1-score (see also [34]). In addition, the absolute displacement between matching true and predicted events was measured. Matching events were defined as those being <600 msec apart from each other. Predicted events lacking a matching ground-truth counterpart were counted as false detections. The *false discovery rate (FDR)* was defined per event type as the number of false detections divided by the number of total event detections. Conversely, true events lacking a matching prediction were counted as false negatives (misses). The false negative rate (FNR) for each event type was defined as the number of missed events divided by the total number of true events.

## 3. Results

Ninety-three minutes of gait activity and 5253 full gait cycles were analyzed across the six patients. The median gait cycle duration ranged from 1045 to 1140 ms, corresponding to cadences between 51 and 59 cycles per minute (see Table 2). The gait cycle duration variability was measured as the median absolute deviation from the median duration and ranged from 10 to 30 ms.

**Table 2.**
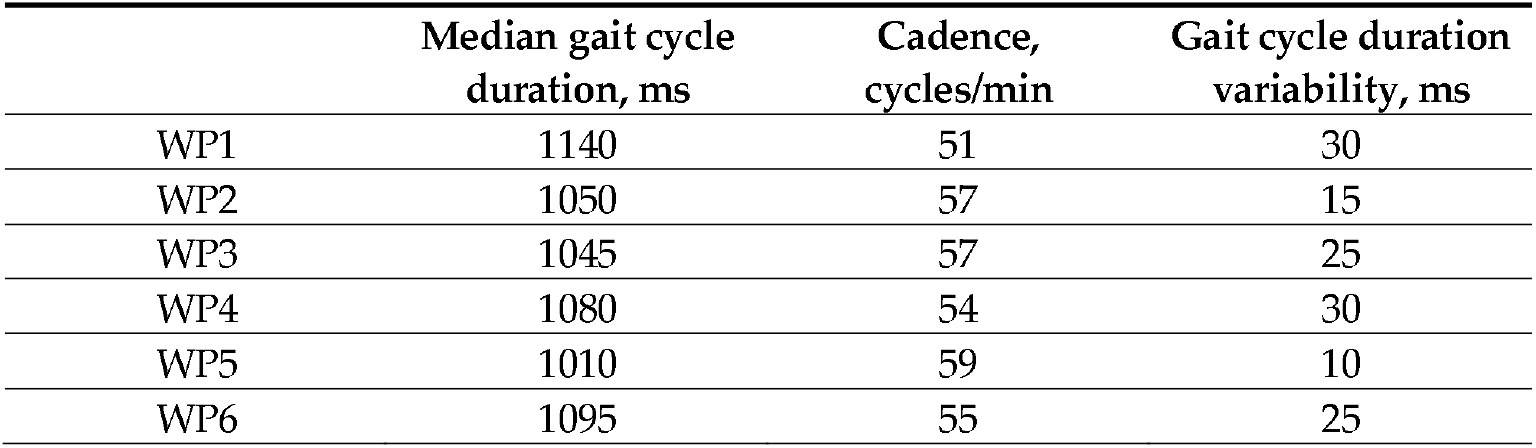
Gait cycle statistics of individual patients.

Figure 2 illustrates the average activation patterns of individual muscles (measured by EMG) relative to the angular velocity profiles (measured by IMU). The upper panels show the average IMU and EMG activity across the gait cycles of all patients as a function of time within a cycle. All ten muscles exhibited stable activation patterns relative to the individual gait events of both legs. Importantly, due to the stable timing of the gait cycle in patients with mild PD, the left leg muscles showed precise activation in well-defined time windows regarding HC and TO events of the left and right leg, and vice versa. The Vl displayed particularly consistent timings (as indicated by dark red colors) both for the left and right leg. The lower panels depict cross-correlations (computed on the concatenated data of all trials) of temporally-shifted EMG activity traces relative to the IMU signal. The same 21 lags were analyzed, ranging from -500 msec to +500 msec relative to the IMU signal reported above for the machine-learning models. Thus, the depicted correlograms represent the independent linear predictive quality of each of the 10*21 = 210 EMG features considered in our models, thereby indicating the influence of each muscle and delay combination for prediction (see also [46]). The activity profiles of all ten individual muscles showed substantial positive and negative correlations with the IMU signal within a window of 1 sec. The highest absolute correlations were observed for the Vl. Specifically, the left Vl activity lagged the left IMU trace by 150 msec (r=0.78) and led the right IMU trace by 350 msec (r=0.76); in contrast, the right Vl activity lagged the right IMU trace by 150 msec (r=0.66) and led the left IMU trace by 350 msec (r=0.67). All reported cross-correlations were statistically significant (p<0.05 after Bonferroni correction).

**Figure 1.**
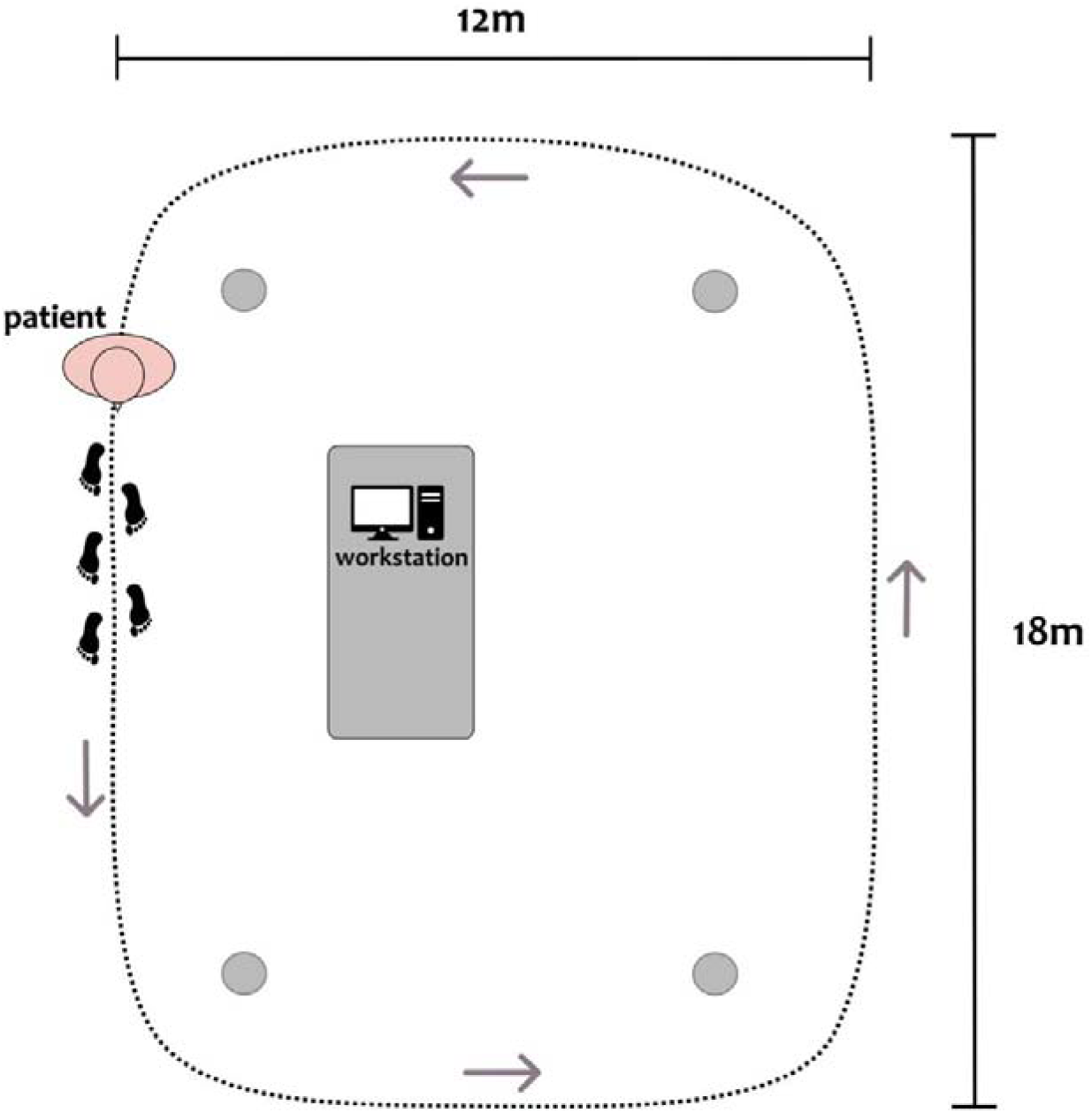
Top-view scheme of the experimental setup, with a patient depicted at the starting position of the circuit. Patients were asked to continuously walk along an elliptical circuit of approximately 60 m around the workstation. The inner boundary of the circuit was marked with four objects at its corners (gray dots). A clinician was close to the patient during all recordings.

**Figure 2.**
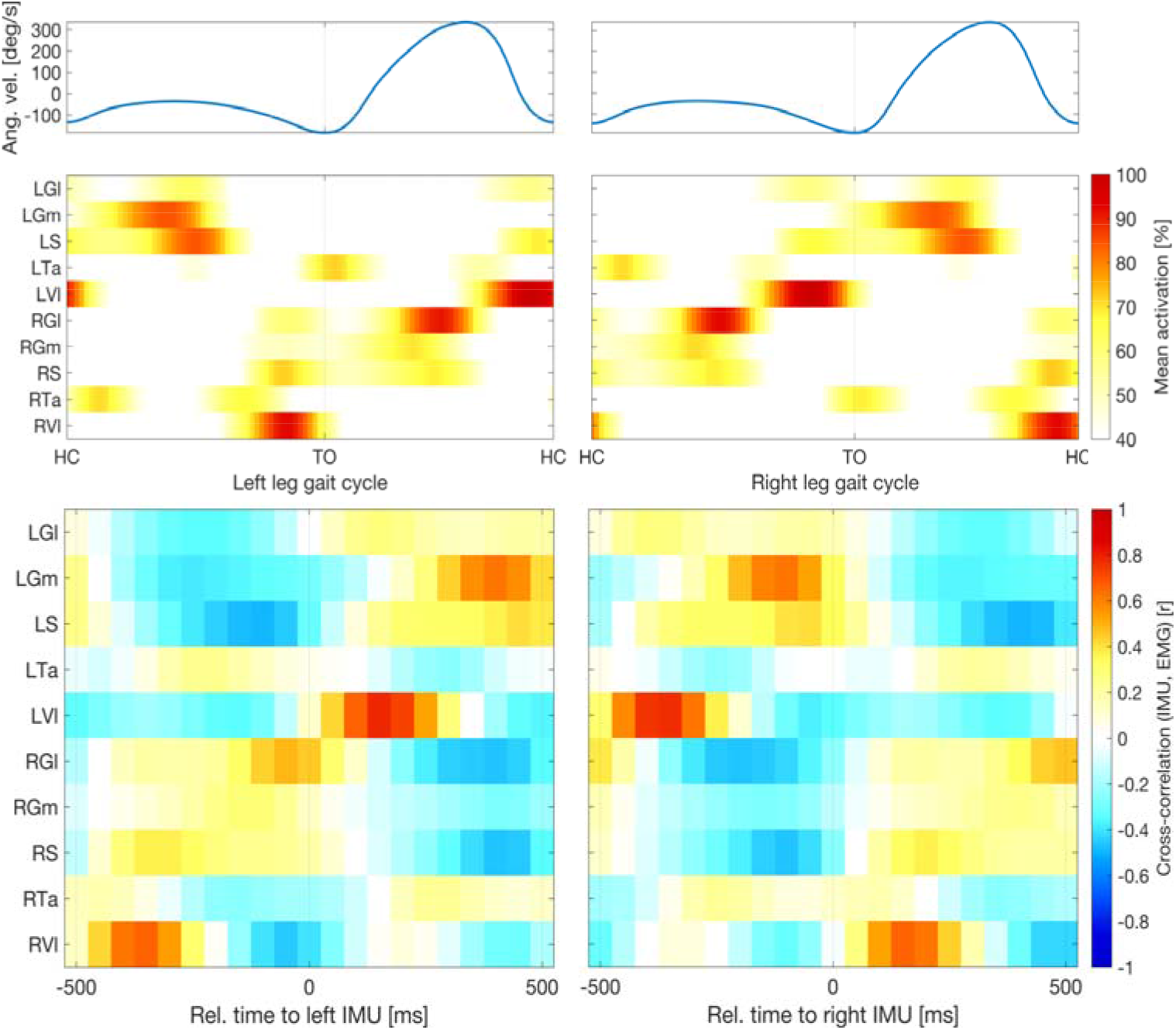
Relative timing of muscular and kinematic signals. Upper panels show average angular velocity measured by inertial measurement units (IMU) and electromyographic (EMG) activity across all gait cycles of all patients as a function of time within cycle. Percentages are relative to the 95th percentile of the raw data. Averages were cropped below 40%. All ten muscles exhibited stable activation patterns relative to the individual gait events of both legs. Lower panels depict cross-correlations (computed on the concatenated data of all trials) of temporally-shifted EMG activity relative to the IMU signal. All ten muscles showed substantial absolute correlations with the IMU signal within a window of 1 sec. Highest correlations (Pearson correlation, r>0.66) were observed for Vl activity at delays of 150 msec relative to the same leg or -350 msec relative to the opposing leg. Abbreviations: left and right gastrocnemius medialis (LGm and RGm) and lateralis (LGl and RGl); left and right soleus (LS, RS); left and right tibialis anterior (LTa, RTa); left and right vastus lateralis (LVl, RVl); TO, toe-off.

Figure 3 shows an example segment of the preprocessed EMG and IMU data of one patient, the EMG-based predictions of the IMU time courses based on all ten available EMG probes, and the gait parameters extracted from true and predicted IMU time series. The EMG time courses of three selected individual muscles (bilateral Ta, S, and Vl) showed the clear periodic pattern of the gait cycle (bottom row). Out-of-sample predictions based on temporal embeddings of the activity of ten muscles showed a high correlation with the true IMU data (top row). Furthermore, gait events extracted from the predicted time series closely matched those extracted from the original IMU traces (top row). True and predicted gait phases based on the extracted events were consequently also closely aligned (center row). Results of similar quality were obtained when predictions were based on the left and right Vl only (see quantitative evaluation below).

**Figure 3.**
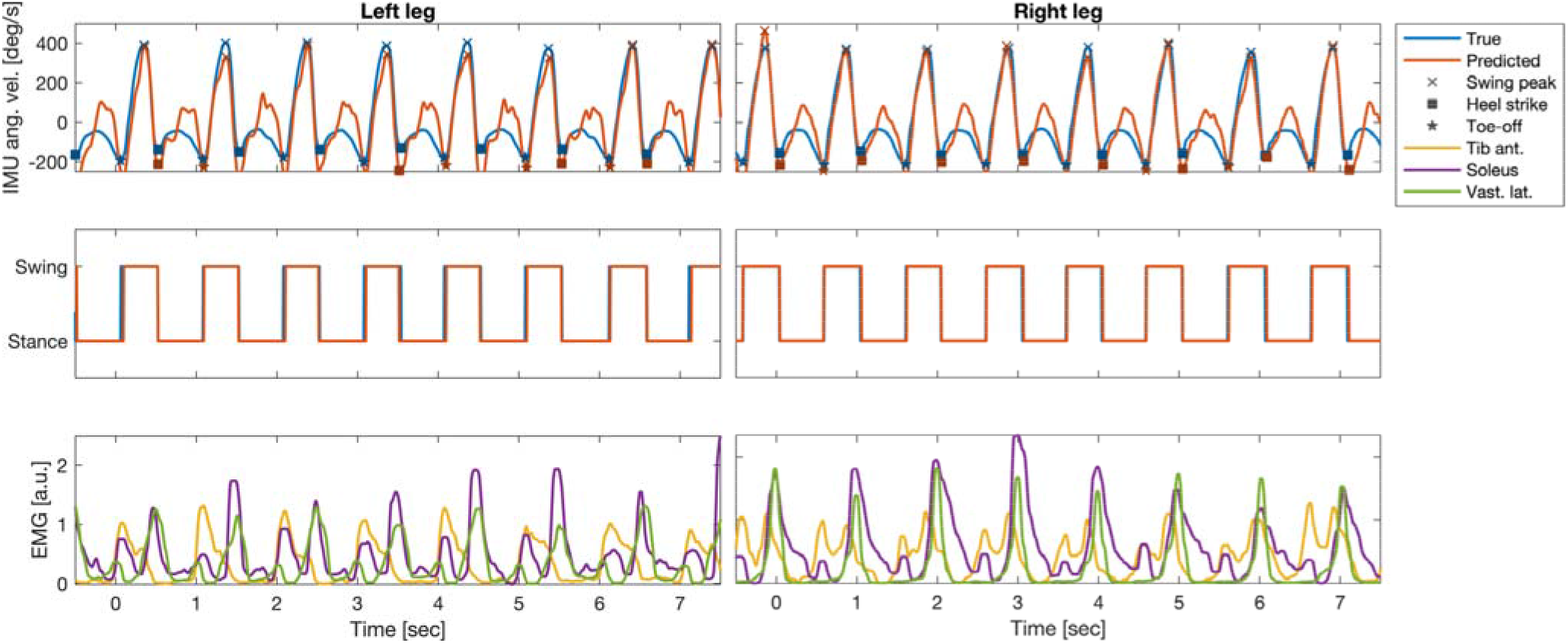
Example segment of the preprocessed electromyography (EMG) and inertial measurement units (IMU); angular velocity at the left and right anklebones) recordings of one patient (P5), as well as the EMG-based predictions of the IMU time courses and the gait parameters extracted from true and predicted IMU time series. Top row: true IMU data and predictions from temporally-embedded EMG activity of ten muscles. Predictions were derived from an ordinary least-squares regression model fitted data of the other five patients. Gait-related events (swing peak velocity, SWP, heel contact, HC, and toe-off, TO) extracted from the predicted time series closely matched those extracted from the original IMU traces. Center row: True and predicted gait phases based on the extracted events were closely aligned. Bottom row: EMG time courses of three selected individual muscles (bilateral soleus, tibialis anterior, and vastus lateralis).

Figure 4 quantitatively summarizes the performance of EMG-based reconstructions of IMU time courses and gait events. The median (IQR across all 26 trials) Pearson correlation between measured and reconstructed IMU time courses, based on all ten muscles, was r=0.80 (0.74 to 0.87) for the left ankle and r=0.85 (0.78 to 0.90) for the right ankle. Using the left and right Vl, the performance was on par, with r=0.86 (0.78 to 0.88) for the left IMU probe and r=0.83 (0.80 to 0.88) for the right IMU probe. Using the left and right Ta and S muscles did not lead to competitive performance, with r=0.47 (0.35 to 0.66) for the left IMU and r=0.55 (0.46 to 0.66) for the right IMU. Importantly, the combination of left and right Vl was found to be on par with the full model for all of performance metrics, whereas the combination of S and Ta was competitive in none. For this reason, we have restricted our reporting to the model comprising left and right Vl. With few exceptions, gait events could be reconstructed with median absolute temporal displacements <50 msec from IMU predictions derived from this model. The median (IQR) displacement for the SWP was 40 (20 to 60) msec for the left leg and 38 (25 to 60) msec for right leg. For HC events, median temporal displacements were 35 (25 to 55) msec for the left leg and 45 (30 to 60) msec for the right leg. For TO events, median displacements were 43 (30 to 100) msec for the left leg and 43 (20 to 95) msec for the right leg. Segmentations of the recordings into dichotomous gait phases based on detected HC and TO events were similar for measured and reconstructed IMU data. Median (IQR) F1 scores were 0.89 (0.87 to 0.93) for the left leg and 0.89 (0.86 to 0.93) for the right leg.

**Figure 4.**
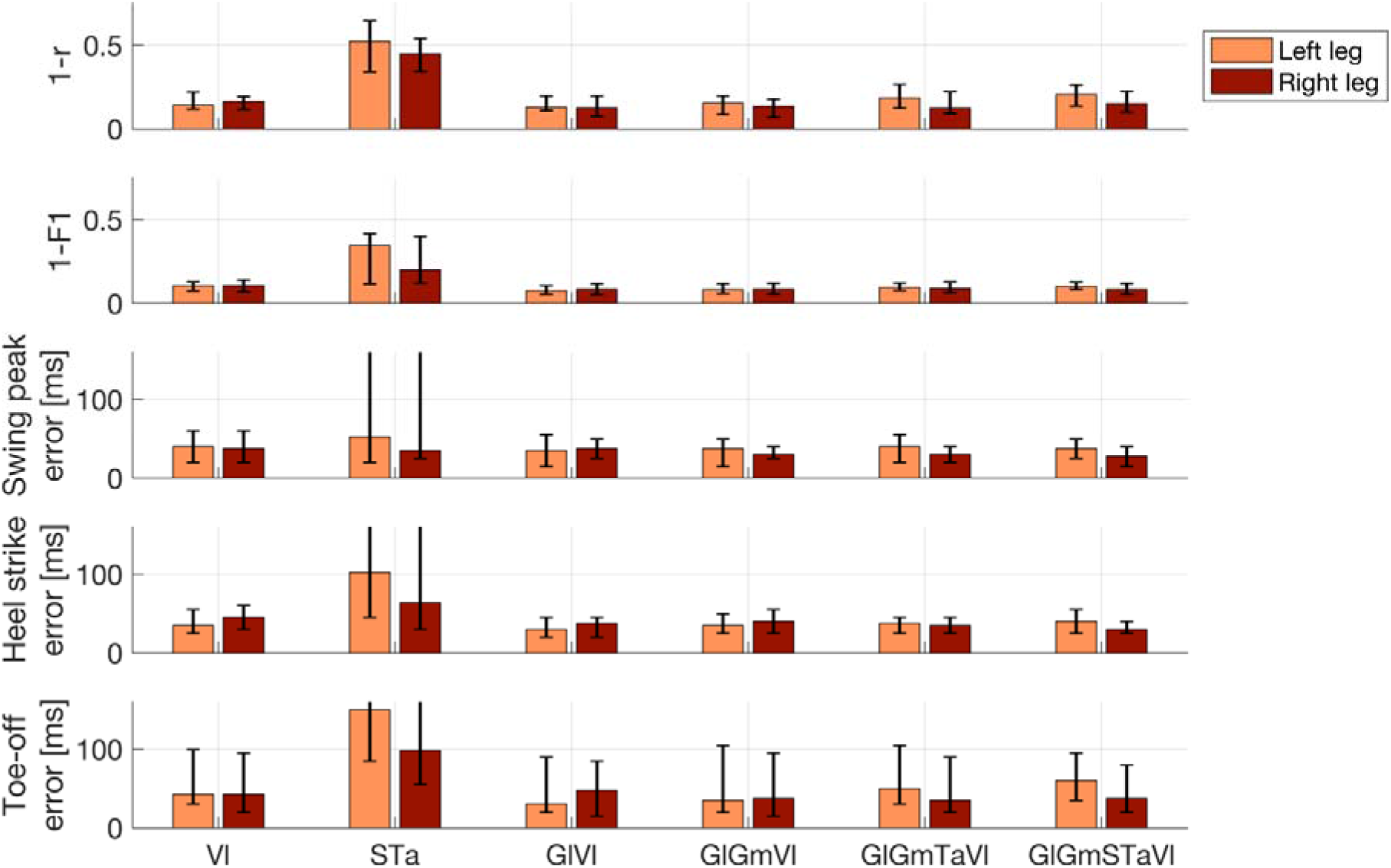
Performance of electromyography (EMG)-based reconstructions of inertial measurement units (IMU) time courses and gait events. Lower numbers represent better performance. Top row: Pearson correlation (r) between measured and reconstructed angular velocity profiles of the left and right ankles. Second row: Accuracy of the reconstructed dichotomous (swing vs. stance) gait phases compared to the IMU-based ground-truth, as measured by the F1 score. Bottom three rows: Absolute displacement of three types of events (swing peak velocity, heel contact, and toe-off) determined from reconstructed rather than measured IMU data. Results are shown separately for the left and right leg and for the best-performing prediction models utilizing between one and five pairs of EMG channels. In addition, results for the combination of the left and right soleus and tibialis anterior are also shown. Bar plots depict median performance across 26 walking trials of six patients in total, while overlaid whiskers depict first and third quartiles. Abbreviations: gastrocnemius medialis (Gm); lateralis (Gl); soleus (S); tibialis anterior (Ta); vastus lateralis (Vl).

Event detection errors were rare and did not occur in most trials. Across all trials, events were missed in 1.4% (n=71; left leg) and 1.3% (n=68; right leg) of cases. Numbers were nearly identical for all three event types, as HC and TO events were always determined relative to the two enclosing SWP events (see Section 2.5). False event discoveries were rare (<0.1% of the total events detected for both legs and all three event types). In absolute terms, between two and four out of over 5000 detected events were false discoveries.

## 4. Discussion

We have demonstrated the feasibility of accurately determining gait events such as HC and TO, defining the swing and stance phases of the gait cycle, in PD patients using a single pair of EMG probes placed bilaterally on the Vl muscle. Our proposed method may have substantial practical benefits in experimental setups in which EMG derivations are indispensable, and where additional equipment for kinematic analysis (e.g., foot switches, IMU, or a motion-capturing system) is either unavailable or would introduce undesired complexity, especially in severely-ill patients. Furthermore, robust acquisition of EMG signals is necessary in experimental and commercial applications to achieve control of myoelectric interfaces for neuroprosthetics [29], including future adaptive DBS devices [30].

Rather than framing the prediction problem as one of binary classification [34], our approach consisted of two steps. First, the angular velocity at the left and right anklebones was predicted from the activity of between two and ten EMG probes. This mapping is learned a priori from training data for which both EMG and IMU recordings are available. Using carefully-designed data features (temporally-embedded, smoothed muscle activation time courses), a simple linear regression approach was found to be suitable to achieve sufficient reconstruction performance. Second, predefined rules were used to extract prominent events and the main phases of the gait cycle. These rules accommodate domain knowledge about the timing of events relative to each other, which constitutes a substantial advantage over algorithms that are completely naïve to the underlying data, framing the gait cycle prediction as an abstract classification problem. Importantly, our approach did not require any calibration involving real IMU data, as models fitted a priori on a training cohort (e.g., the data reported here) can be readily applied to new patients. Due to the simplicity of our model, its application amounts to a simple linear filtering of the appropriately recorded and preprocessed EMG data and does not require any advanced machine-learning software. In addition, our approach of approximating IMU time courses instead of individual events or categorial segmentation labels offers numerous additional advantages. These include direct interpretation of the predicted time courses in terms of gait mechanics. Potential failure modes of the model (e.g., due to misplaced or noisy EMG probes) could easily be detected through visual inspection of the predicted time courses. Since the SWP could be accurately detected even from reconstructed angular velocities, and HC and TO were defined relative to SWP, our system achieved low numbers of event-detection errors and a high overall accuracy regarding the determination of gait phases. It is also likely that our approach could be generalized to the extraction of other biomechanically-relevant parameters of the upper and lower extremities.

Contrary to our prediction, the EMG profiles of the S and Ta muscles were insufficient to reliably identify major gait cycle events in parkinsonian patients. We based this hypothesis on the distinctive and synergistic activity of these two monoarticular (i.e., ankle) muscles during human locomotion. Indeed, normal EMG activity for the plantar flexors has been reported to occur mainly during the stance phase. In this phase, the triceps surae restrains tibial rotation controlling for the disequilibrium torque, which is responsible for propelling the body [47,48]. The ankle dorsi-flexors are instead mainly active during the swing phase, controlling for sufficient foot clearance, with an additional contribution in the loading response phase for the lowering of the foot to the ground after HC [49], thus assisting the forward momentum of the tibia during the heel rocker action at the ankle [50]. These muscles, however, may show large stride-to-stride variability in the EMG profiles [49], especially in patients with PD [51,52]. In particular, a great intra- and inter-subject variability of the Ta activity during gait has been described in parkinsonian patients in the meds-off state [51].

The prediction model did not improve when replacing the S muscle with the Gm or Gl, or by adding this muscle to the S-Ta pair (data not shown). This was unexpected because while the S muscle may provide less forward propulsion with physiological aging, the gastrocnemius muscle has been shown to maintain its contribution in initiating swinging limb movement [53,54], thus possibly allowing a more accurate detection of kinematic events. Rodriguez and colleagues demonstrated a simplification of modular control of locomotion in PD with an individual muscle contribution of the gastrocnemius, but not the S, among ankle plantar flexors and the semimembranosus and biceps femoris for the knee flexor musculature [55].

In our study, EMG recordings of the Vl provided the most accurate prediction of IMU times series and gait events. The action pattern of this muscle during the gait cycle paralleled the activation of the Ta, but was more selectively confined to the HC. This muscle controls the knee flexion that occurs after HC and ensures knee extension during terminal swing to prepare for ground contact [50,56].

In principle, there are an infinite number of different combinations of muscle activations that can be applied to maintain a particular posture or produce a given movement [57]. However, despite the apparent redundancy, four or five component activity patterns may be distributed to all the muscles that are specifically activated during locomotion; thus, the activation of each muscle involves a dynamic weighting of these basic patterns [58,59]. Interestingly, Ta, S, and Vl contributed differently to these factors [58,59]. Our results suggest that characteristic activity patterns of one pair – the left and right Vl – are sufficient for proper detection of gait events in patients with PD (H&Y: I-III).

### 4.1 Limitations

Our study is somewhat limited by the fact that IMU data are not considered the “gold standard” for defining ground-truth gait parameters. Force plates would have allowed precise detection of HC and TO events, and possibly the individual muscle contribution to ground reaction forces [60,61]. However, it would have been impracticable to record the high number of steps and total gait time acquired in our study using force plates. IMU systems are sufficiently accurate in the assessment of fundamental gait spatiotemporal parameters [23,62] and have previously been used as ground truth for gait event detection [63]. Furthermore, they allow detection of the SWP event, which would not have been captured by ground devices, foot switches, or insole pressure sensors.

The proposed approach was not tested on healthy control data. However, we expect our model to effectively predict gait events in healthy controls as patient data are more heterogeneous and generally more challenging in terms of gait alterations, inter-subject, and inter-trial variability, as well as artifact contamination. We were also only able to recruit a few patients for this study. However, it should be considered that walking for over three minutes in the meds-off state is very challenging for subjects with PD and greatly limited patient recruitment. Another limitation was the relatively homogeneous walking speed across all patients. We preferred not to alter the patients’ natural speed because we wanted to test our model in an ecological setup. Also, the meds-off state limited the recording window and the possibility of exploring more than one gait condition. It is thus presently unclear how well our prediction model would perform for different speeds when applied out-of-the-box. However, it is straightforward to adapt the model to different speeds by either temporally adjusting the embedding delays τ, …, τ_*k*_ of test participants to their individual walking speed or retraining the model on data with matching speed.

## 5. Conclusions

We have demonstrated the accurate and robust detection of gait events in six parkinsonian patients using just two EMG probes placed on the left and right vastus lateralis. Unlike solutions presented in previous work, our approach proceeds in two steps. First, IMU time courses are predicted from EMG activity within a surrounding temporal window using multiple linear regressions. And, second, gait parameters such as heel strikes and toe-off events are extracted from the predicted time series. This approach led to accurate results and has the advantage over previous ones that discrete gait events and continuous time series of relevant kinematic quantities can be predicted. It is further expected to generalize to the extraction of further gait parameters not considered here without any model retraining. Our model, an example dataset, as well as Matlab code for data preprocessing, model training, model evaluation, and plotting, is made publicly available^1^. Our approach may have practical benefits for gait studies in which the application of multiple sensing devices is considered impractical, troublesome, or too expensive. Notably, our model was validated using a leave-one-patient-out strategy. We observed very good performance on held-out patients, demonstrating that the model is able to accommodate the across-patient variability of the studied clinical population. Future work will adapt our approach to varying walking speeds and may further extend it to the prediction of other kinematic data obtained from EMG.

## Data Availability

The data presented in this study are available upon request from the corresponding author. The data are not publicly available for privacy reasons.

## Author Contributions

Conceptualization, S.H. and I.U.I; methodology, S.H., C.P. and I.U.I.; software, S.H.; formal analysis, S.H. and F.P.; investigation, C.P. and I.U.I.; resources, S.H., C.P. and I.U.I.; data curation, C.P.; writing—original draft preparation, S.H. and I.U.I..; writing—review and editing, S.H., C.P., F.P., and I.U.I.; visualization, S.H. and C.P.; supervision, C.P.; project administration, I.U.I.; funding acquisition, S.H., C.P. and I.U.I. All authors have read and agreed to the published version of the manuscript.

## Funding

This study was sponsored by the Deutsche Forschungsgemeinschaft (DFG, German Research Foundation) – Project-ID 424778381-TRR 295 and the Fondazione Grigioni per il Morbo di Parkinson. SH received funding from the European Research Council (ERC) under the European Union’s Horizon 2020 research and innovation program (Grant agreement No. 758985). CP was supported by a grant from the German Excellence Initiative to the Graduate School of Life Sciences, University of Würzburg. IUI received funding from the New York University School of Medicine and The Marlene and Paolo Fresco Institute for Parkinson’s and Movement Disorders, which was made possible with support from Marlene and Paolo Fresco.

Institutional Review Board Statement: The study was conducted in accordance with the Declaration of Helsinki and approved by the Ethics Committee of the University of Würzburg (n. 103/20 and 36/17).

## Informed Consent Statement

Informed consent was obtained from all subjects involved in the study.

## Acknowledgments

We would like to thank all patients and caregivers for their participation. The draft manuscript was edited for English language by Deborah Nock (Medical WriteAway, Norwich, UK).

## Conflicts of Interest

The authors declare no conflict of interest. The funders had no role in the design of the study; in the collection, analyses, or interpretation of data; in the writing of the manuscript; or in the decision to publish the results.

## Footnotes

1 https://github.com/braindatalab/EMGgaitprediction

## Notes

### Competing Interest Statement

The authors have declared no competing interest.

### Author Declarations

The Ethics Committee of the University of Wuerzburg gave ethical approval for this work.

